# Cortical Manifolds in Cognitive Recovery following Supratentorial Neurosurgery

**DOI:** 10.1101/2025.01.16.25320459

**Authors:** A Poologaindran, R Romero-Garcia, AI Luppi, MG Hart, T Santarius, S Price, ME Sughrue, RAI Bethlehem, S Sonkusare, Y Erez, J Suckling

**Author notes:** **Corresponding Authors** Anujan Poologaindran, PhD, Brain Mapping Unit, Department of Psychiatry, University of Cambridge, Professor John Suckling, Brain Mapping Unit, Department of Psychiatry, University of Cambridge.

## Abstract

The cerebral cortex is topographically organized to integrate and segregate unimodal (e.g. sensorimotor) and transmodal brain networks to scaffold cognition. Cortical gradient mapping provides a framework to examine the relationship between connectivity patterns of macroscale functional brain networks within a low-dimensional (manifold) space. Using this technique, we longitudinally examine how diffuse gliomas, their neurosurgical resection, and subsequent cognitive rehabilitation impact the topographic organization of brain networks. First, using UKBioBank data (n=4000), we validate the general assembly of cortical gradients in healthy individuals. Next, using CamCan data (n=620), we found that gradient dispersion relates to executive functions (EFs) across the lifespan. Finally, in diffuse glioma patients undergoing neurosurgery (n=17, 59 unique scans), we observed that gliomas integrate into the cortical manifold by reducing gradient dispersion compared to healthy controls. This finding was replicated in an independent cohort and contrasted with meningioma patients. Finally, long-term cognitive improvement after surgery was linked to increases in gradient dispersion, while long-term deficits were associated with decreases in gradient dispersion; longitudinal analyses revealed month 3 (not month 12) as the crucial window for gradient reconstitution. Overall, diffuse gliomas minimally disrupt the assembly of cortical manifolds, but the ability to reorganize the cortical manifold within 3 months post-surgery is predictive of long-term cognitive outcomes. By investigating neurosurgical patients with atypical neuroanatomy, this study contributes to the expanding literature on how aging, disease, and pharmacological interventions impact cortical gradients. Future studies are warranted to assess the utility of mapping cortical manifolds in neurosurgical patients.

## Introduction

Low-grade gliomas (LGGs) are slow-growing infiltrative brain cancers that embed themselves within the brain’s structural connectome^1^. Unlike strokes, which acutely damage the brain’s connectome, LGGs gradually and dynamically remodel it in more insidious ways^1,2^. Recent seminal investigations have demonstrated that neurons and glioma cancer cells form functional synapses, and excitatory glutamatergic neurotransmission drives tumour progression^3,4^. Moreover, evidence from human neuroimaging demonstrates that gliomas preferentially infiltrate associative cortical areas which contain distinct connector hubs, stem-like cells, and transcription factors for gliomagenesis^5^. Supramaximal resection of LGGs can confer prolonged progression-free survival, reduced propensity for malignant transformation, and increased overall survival regardless of the glioma’s molecular subtype^6-9^. Nevertheless, LGG patients often have unpredictable post-operative cognitive trajectories; nearly 50% of patients experience some cognitive difficulties in language, memory, and/or executive functions^10-11^, which may further be exacerbated by radiotherapy-induced cognitive deficits^12^. Nearly 25% of LGG patients who experience cognitive deficits report lower rates of return to full-time work, strained personal relationships, and reduced independence^13^. Ideally, throughout neuro-oncological care, ‘onco-functional’ balance, that is maximizing extent of resection while minimizing neurological deficits, should be prioritized by assessing the state of functional brain networks governing cognition and informing the timing and approach to cognitive rehabilitation strategies^14^.

Human neuroscience is embracing a shift away from the ‘localizationist’ perspective of brain function towards a network-based perspective positioning the brain as a coordinated, complex, inter-connected network-of-networks^15,16^. The emergence of higher-order transmodal cortical areas results from the convergence of information across unimodal sensory and motor areas^17^. This suggests a measurable processing gradient for understanding the relation between domain-general and domain-specific cortical areas^18^. By characterizing the placement of cortical areas within a broader cortical hierarchy, we can form the foundation to better characterize high-dimensional neural data into a single low-dimensional space^19^. Furthermore, the ability to describe brain-wide organizational principles as a manifold offers an analytical paradigm to investigate how distinct brain systems integrate and to give rise to higher-order cognition.

To date, no studies have longitudinally assessed the impact of brain cancer, neurosurgery, or cognitive recovery on the hierarchical organization of brain networks (herein referred to as cortical manifolds). ^14,20^. While previous studies have leveraged gradient mapping to describe atypical gradient organization in schizophrenia^21^, autism^22^, psychedelic use^23^, consciousness^42,43^, ageing^24^, applying this technique in the context of cancer, neurosurgery, and cognitive rehabilitation is a step towards clinically-actionable connectomics given the disease and treatment goals of preserving onco-functional balance^25,26^. Thus, we aimed to utilize state-of-the-art gradient approaches to explore how cortical gradients are affected by gliomas, neurosurgery, and rehabilitation. We hypothesized that the degree of manifold tethering may stratify cognitive functioning in both healthy individuals and glioma patients undergoing neurosurgical intervention.

## Methods

To begin with, the UKBiobank dataset of n=4000 healthy individuals with functional connectivity data was harnessed to validate the general assembly of cortical gradients. Next, the CamCAN dataset comprising of 620 healthy individuals with ages across the lifespan was utilized to determine how dispersion of cortical gradients relates to executive functioning. Using this fundamental insight, we probed how cancer, surgery, and rehabilitation affected cortical gradients and cognitive outcomes in a prospective, longitudinal, clinical trial of patients undergoing glioma surgery. Overall, our goal was to determine if clear relationships existed between the assembly and dispersion of cortical gradients and cognitive functioning in both health and disease.

**Figure 1A/B.**
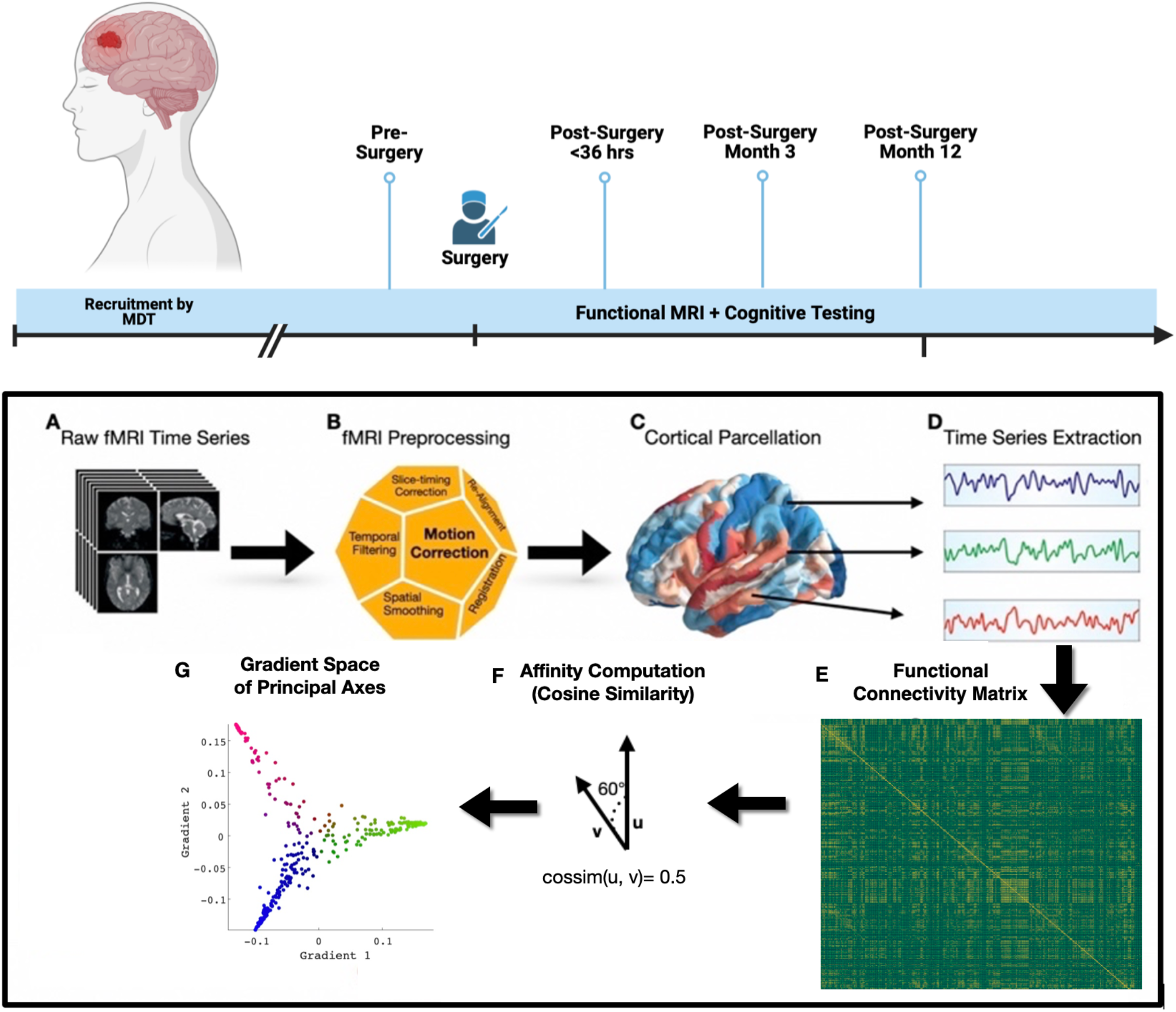
Neurosurgical Study Design and Functional Gradient Mapping Workflow. **Top Panel:** Participants were recruited by a multi-disciplinary neuro-oncology team and if they were eligible for surgical resection. Data collection included resting-state functional MRI (rs-fMRI) and cognitive assessment at multiple time points: pre-surgery, immediate post-surgery (<72 hrs), and at months 3 and 12 post-surgery. **Bottom Panel:** A: rs-fMRI was acquired from all participants at each time point. B: Data underwent pre-processing steps to accountfor head motion. C: After being registered to anatomical space, each patients’ cortex was parcellated by the Glasser HCP scheme comprising of 360 regions. D. For each parcel, the BOLD time series was extracted. E. Time series from all parcels were correlated with one another to estimate functional connectivity (FC). F. The affinity matrix was then calculated from the original FC for each rs-fMRI acquisition. G. Diffusion embedding was applied to produce a low-dimensional manifold of the FC data. H. Final connectivity matrix after sorting.

### MRI Pre-Processing, Lesion Masking, and Functional Time Series Extraction

Details of structural MRI processing, lesion marking, resting-state functional MRI (rsfMRI) pre-processing, and lesion masking have been previously reported^26^. Importantly, the Glasser parcellation scheme from the Human Connectome Project (HCP) was chosen to parcellate the cortex into 360 defined regions to establish a common nomenclature with other neurosurgical connectomic studies^28-30^. Moreover, previous methodological work demonstrated that utilizing <200 nodes may not reliably capture putative functional boundaries and greater consistency is achieved when using >300 anatomically-informed nodes^19^.

### Functional Gradient Mapping

We computed cortical gradients derived from resting-state functional MRI functional connectivity (FC) represented as weighted whole-brain graphs. Cortical gradients are defined as the axes of principal variance in cortico-cortical FC mapped nonlinearly onto a low-dimensional manifold^18,19,22^ and were created with the Brainspace toolbox^19^. Since we were interested in the ensemble relationships between cortical regions, consistent with previous studies^18,22^ and default parameters^19^, the cosine similarity kernel function was calculated generating an affinity matrix which captures the node-wise similarity of FC by computing the cosine of the angle between the corresponding feature vectors. Cosine similarity ranges from -1 to 1, and negative correlations were transformed to non-negative values using the normalized angle kernel. To identify the ‘ordering’ of the input matrix in a lower dimensional manifold space, the affinity matrix underwent diffusion map embedding (DME), a non-linear dimensionality reduction that decomposes the matrix into a set of principal eigenvectors describing axes of maximal variance^31^. DME was employed to resolve the gradients of subject-level connectomes. Given that FC data contain both local- and long-range connections, DME translates these relationships into distances and represent the global connectivity structure as a distribution of cortical points in an embedding space^31^. Cortical points that are strongly connected by either many weak connections or few very strong connections are close in this space, whereas points without connections are far apart^18^. Consistent with previous studies, ten components were generated based on Brainspace parameters set to sparsity 90, alpha 0.5, and diffusion time zero^18,19,21,22^. For each participant’s matrix and group-averaged matrix, the scree plot was checked to ensure a minimum between the first two components and the rest of the remaining eight components. To visualize the data, the first and second principal components were plotted.

### Centrography, Procrustes, and Statistical Analyses

Bethlehem and colleagues demonstrated that between- and within-network gradient dispersion in frontoparietal, attentional, and default mode network was negatively associated with cognition^24^. Thus, to quantify overall cortical gradient dispersion, we utilized centrography analysis of point pattern analyses^32^. In brief, the standard distance deviation was calculated for each cortical gradient map in two dimensions. The *Standard Distance (SD)*, also known as the *Standard Distance Deviation*, is the average distance by which all points vary from the *mean centre*, measuring the compactness of a distribution. It is visualised as the radius from the *mean centre* of a circle plotted as an indication of dispersion. The SD is calculated as:

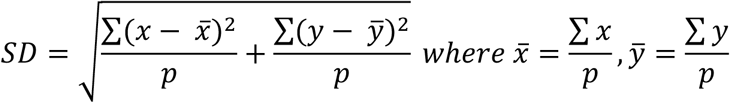

where *p* is the number of points. To compare the shapes of gradients, Procrustes statistical shape analyses compares two similarity matrices while ignoring the effects of translation, scaling, and rotation^36^. Non-parametric t-tests were utilized to compare SD cross-sectionally between patient and control samples, and ANOVA to test the SD between surgical participants with long-term improvements and long-term deficits.

### Factors Influencing Gradients Dispersion

To identify which brain regions (and networks) contribute to maximizing or minimizing gradient dispersion, we determined which brain regions that maximized the SD formula for each subject. Specifically, the numerator in both terms of the SD formula calculates how much a given brain region’s gradient position deviates from the individual’s overall average, with larger deviations indicating a greater distance. By comparing these distances across different brain regions, we aimed to determine whether one or more brain networks was consistently involved in either maximizing or minimizing gradient dispersion. A chi-square goodness-of-fit test was used to evaluate whether certain brain regions were involved more often than expected under a uniform distribution model.

### UKBioBank Dataset

Initially, the general organising principle of functional gradients was validated in 4000 healthy individuals from the UKBiobank^33^ from whom an anatomical structural T1-weighted MRI and resting-state functional MRI (rsfMRI) scans were available. Details of participant selection, imaging acquisition parameters and processing have been reported previously^33^. For the entire cohort, a group-averaged functional connectivity matrix was constructed and processed with the cortical gradient mapping workflow. The shape and assembly of the cortical gradients in the first two principal components were then qualitatively compared to extant literature reporting on cortical gradients also constructed from rsfMRI in independent datasets^18,19,22-24^.

### cam Dataset

Data from healthy participants across the lifespan were acquired as part of the Cambridge Centre for Aging and Neuroscience (Cam-CAN) dataset which comprised of healthy individuals aged 18-88 years. The study has been described in detail elsewhere^34^. Cam-CAN (n=620) participants and our surgical cohort (described below) were scanned on the same Siemens Prisma 3T MRI scanner with the same acquisition parameters. Thus, Cam-CAN participants were propensity-score matched to our surgical cohort in a 1:10 ratio for age (+/-1) and sex and formed the control sample. To probe how cortical gradient dispersion related to higher-order cognition, each control participant’s cortical gradients were calculated and the resulting SD estimates were then related to the Cattell fluid intelligence (total) score, which measures executive functioning and the ability to solve novel reasoning problems^35^.

### CAESAR Experimental Medicine Study

In the CAESAR prospective, longitudinal, experimental medicine study of patients undergoing LGG surgery, participants were recruited by the adult neuro-oncology multidisciplinary team at Addenbrooke’s hospital (Cambridge, UK). The study was approved by the Cambridge Central Research Ethics Committee (Reference number 16/EE/0151) and all patients provided written informed consent. Participant characteristics, inclusion and exclusion criteria, and image scanning parameters have been previously described^27^. Functional connectomic data was acquired pre-surgery, postoperative Day 1, and 3 and 12 months in the rehabilitation period; four time points. In total, the following neuroimaging data in patients was collected longitudinally: n=17 (pre-surgery), n=17 (post-surgery Day 1), n=13 (post-surgery Month 3), and n=11. Thus, n=11 patients had a full dataset of neuroimaging data available at all timepoints. Table 1 in the Supplementary Material lists which patients underwent neuroimaging at each time point.

**Table 1.**
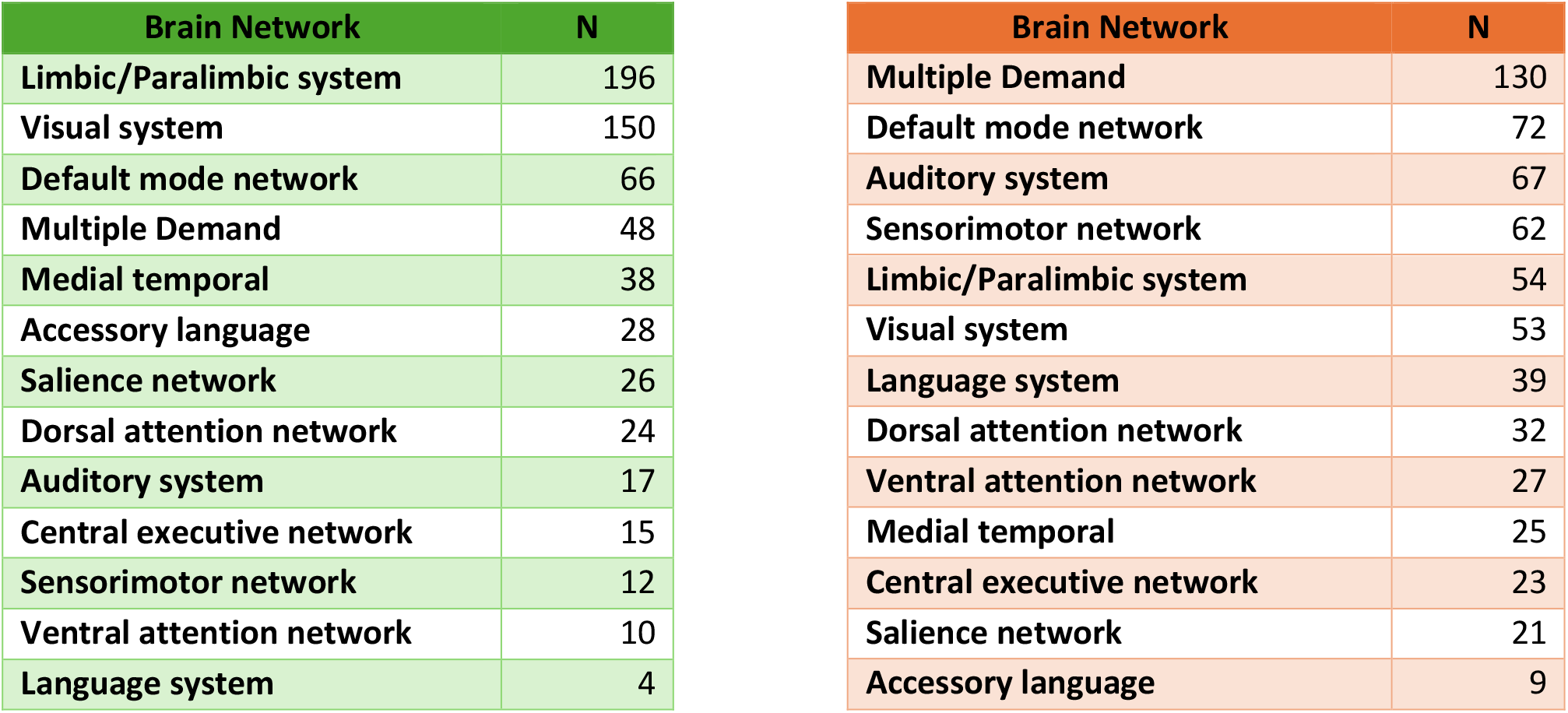
Brain Networks Maximizing (Left) and Minimizing (Right) Gradient Dispersion. N indicates number of subjects in which a brain region’s corresponding network was consistently involved in gradient maximization (left) or minimization (right)

Details of neuropsychological assessment has been previously reported^36^. Patients were stratified as either long-term ‘improvers’ if they improved or did not deteriorate from pre-surgical cognitive functioning following surgery, or ‘decliners’ if they deteriorated from pre-surgical cognitive functioning following surgery. Differences between improvers and decliners in cortical gradient dispersion was assessed with non-parametric t-tests and spearman’s rank correlation. This demarcation between ‘improvers’ and ‘decliners’ informed all subsequent neuroimaging analyses.

## Results

### General Assembly of Normative Cortical Gradients

Individual connectivity matrices derived from the UKBiobank resting-state functional MRI data (n=4000) were aggregated to create a group-averaged matrix (Figure 1A). Following DME, the first two principal components explained 48% of the data (scree plot; Figure 1B). Plotting the first two PCs resulted in a principal gradient of intrinsic FC that placed transmodal associative cortical areas (green points) at a maximal distance from primary regions specialized in sensorimotor (red points) or visual (blue points) functions; Figure 1C. This general assembly of cortical gradients closely resembles previous work^18, 19, 21-23^.

**Figure 1.**
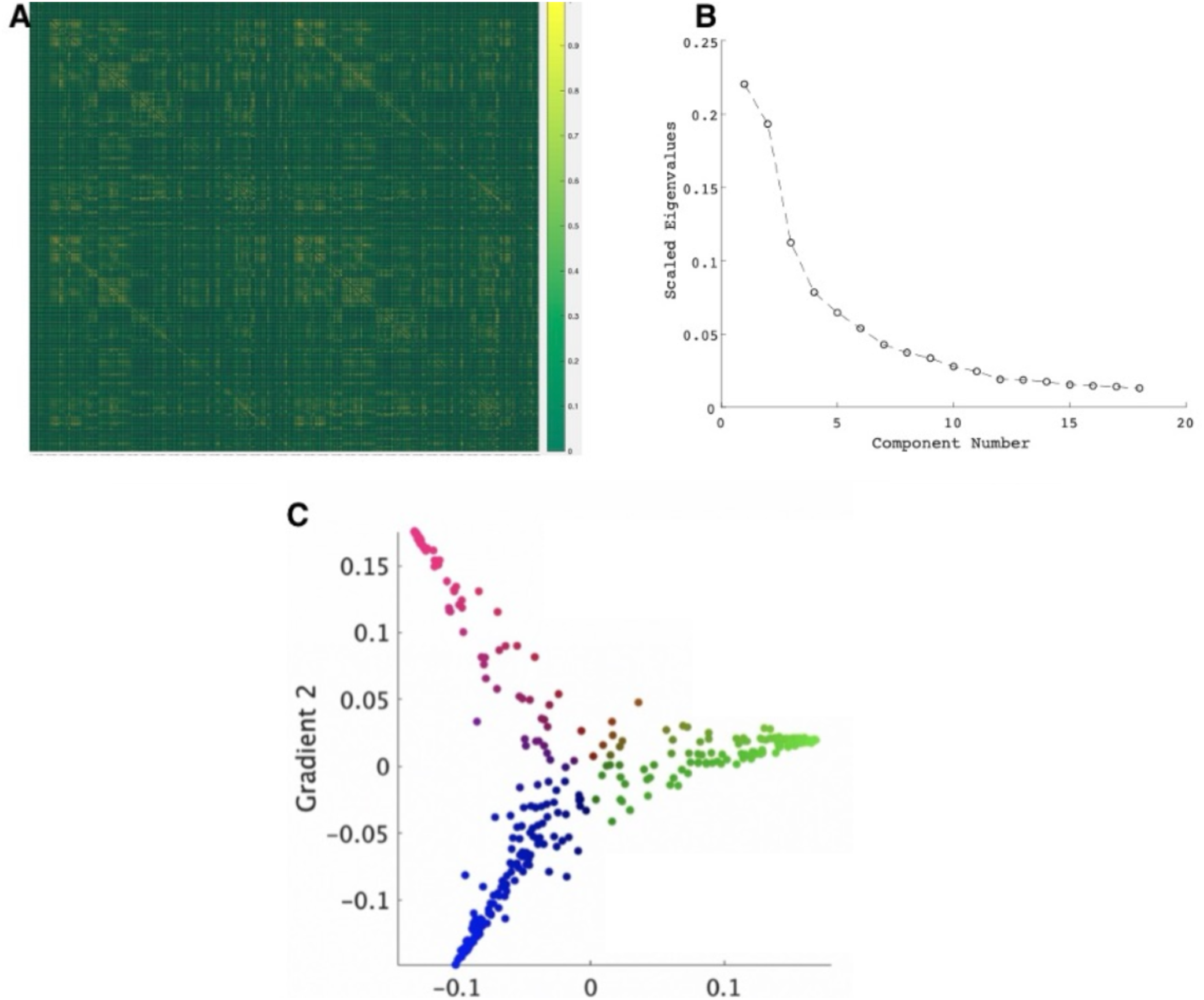
UKBiobank (n=4000) Principal Gradients of Macroscale Cortical Organization. **Caption Figure 1**: A) Group-averaged UKBioBank functional connectivity matrix B) Screeplot with first two principal components. C) Cortical gradient hieararchy of processing from unimodal (blue-visual, red-sensorimotor) to transmodal-(green-associative) areas as initially proposed by Mesulam (1998) and formalized by Margulies et al. (2016). This figure was produced using the BrainSpace toolbox^19^.

### Cortical gradients are related to executive functioning

The group-averaged connectivity matrix from the CamCAN dataset (n=620) was processed by dimensionality reduction with the first two principal components explaining 43% variance in the data. For each participant, the first two PCs were plotted (Figure 2A) and the SD (Figure 2B) calculated to measure gradient dispersion. Multiple linear regression (Figure 2C), while controlling for age, sex, and age-by-sex interactions, revealed a significant and moderate negative relationship between gradient dispersion and executive functioning measured by Cattell fluid intelligence (R^2^=0.4534, p=2.2×10^−16^, effect of dispersion in model: slope=-12.11, std=4.98, p=0.0153; effect of age in model: slope= -0.24677, std=0.011, p= <2e-16; effect of Male gender in model: slope= 0.94487, std=0.408, p=0.0210). Thus, highlighting a more tethered cortical hierarchy is related to improved performance on executive functioning.

**Figure 2.**
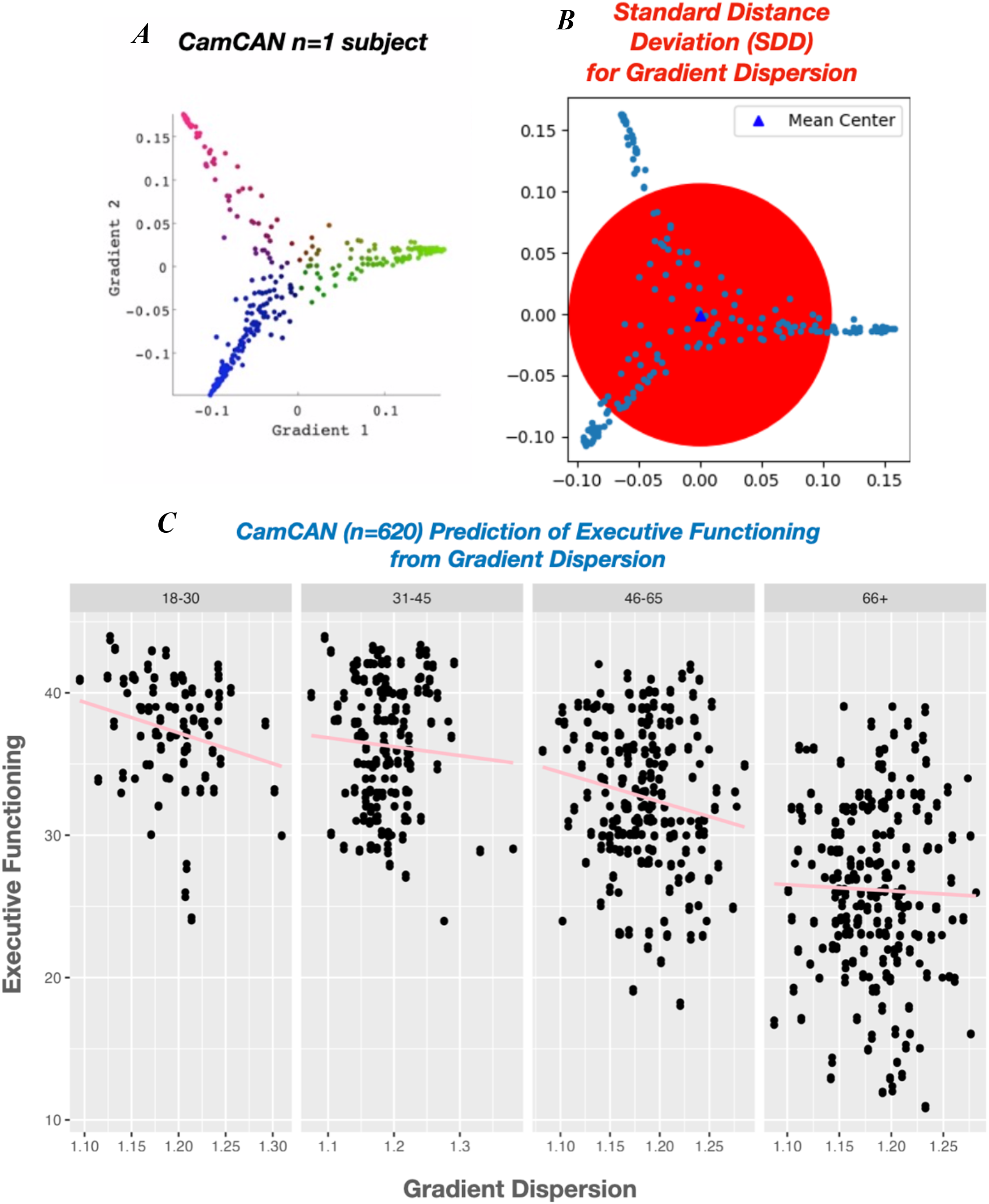
Relationship between Dispersion of Cortical Hierarchy and Executive Functioning in Healthy Individuals. Figure 2: A) An example single-subject (CamCAN) cortical gradient. B) For each participant in dataset (n=620), the standard distance (SD; radius of red circle) was calculated to quantify gradient dispersion within the space of the first two principal components. C) Multiple linear regression, adjusting for for age, sex, and an age-by-sex interaction, was performed to predict executive functioning scores from gradient dispersion indices, and a significant negative relationship detected (complete model; R^2^= 0.4534, p=2.2×10^−16^; effect of dispersion in model; slope=-12.11, std=4.98, p=0.0153). Scale on y-axis refers to overal Cattell scores for executive functioning.

### Brain Network and Biological Drivers of Gradients Dispersion

When evaluating which brain regions were consistently involved in maximizing gradient dispersion, we found that regions within the limbic/paralimbic network, followed by the visual network, played a key role in maximizing SD (Table 1). Conversely, regions responsible for minimizing gradient dispersion were primarily part of the Multiple Demand system, followed by the Default Mode network, and were involved in reducing SD. Table 1 summarizes the regions associated with the maximization and minimization of SD. Chi square goodness-of-fit results for maximization and minimization of gradient dispersion.

### Gliomas alter cortical gradient dispersion

The glioma distribution of the CAESAR cohort (n=17) was frontotemporally dominant; Figure 3A. The group-averaged pre-surgical connectivity matrix was processed by DME with the first two principal components explaining 39% of the variance; Figure 3B. A non-parametric t-test of the SD derived from individual cortical gradients between pre-surgical patients and control participants revealed non-overlapping values and thus a highly significant difference (p=6.9×10^−7^); Figure 3D. Statistical shape analyses via Procrustes analysis revealed non-significant differences between glioma patient gradients (Figure 3C) and normative control gradients (Figure 3E), indicating a maintenance of gradient assembly but significant glioma-induced dispersion differences. Comparison between glioma and healthy control gradient dispersion was replicated in an independent LGG (Figure S1) cohort and contrasted against meningioma patients who did not significantly differ with control participants on SD (Figure 4).

**Figure 3.**
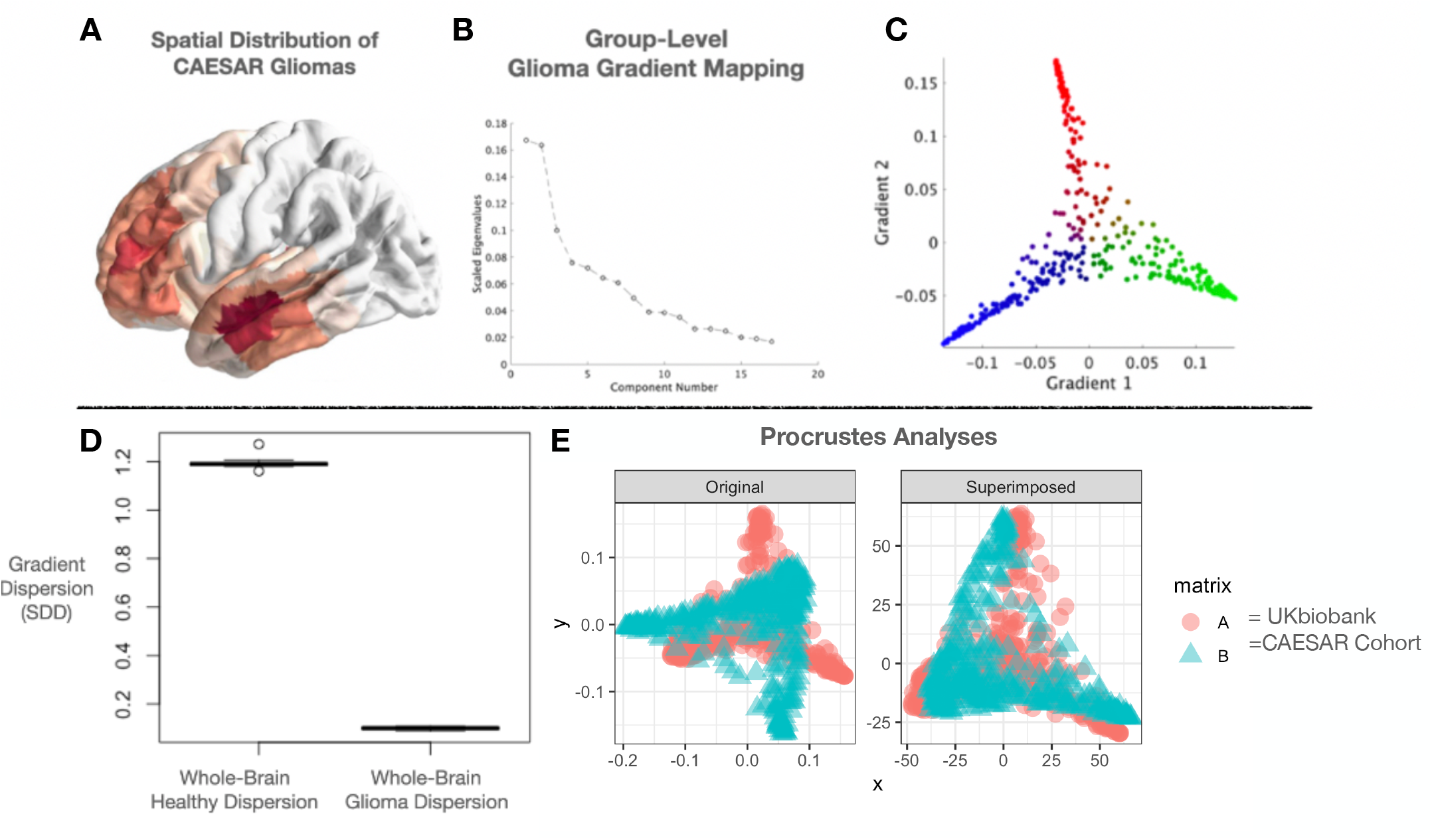
Gliomas Intricately Integrate within Hierarchical Brain Networks. **Top Panel A**. Spatial distribution of gliomas plotted on the cortical surface. **B**. Screeplot with first two principal components of the group-averaged connectivity matix derived from pre-surgical rs-fMRI. **C**. Despite glioma presence, cortical gradients remain largely unperturbed. **D**. Significant decrease in the standard distance (i.e. gradient dispersion) due to glioma compared to healthy controls (Cohen’s d=66.4, magnitude=large, p=6.9×10-7). **E**. Procrustes shape analysis revealed no significant difference in shape between glioma gradients and control gradients.

**Figure 4.**
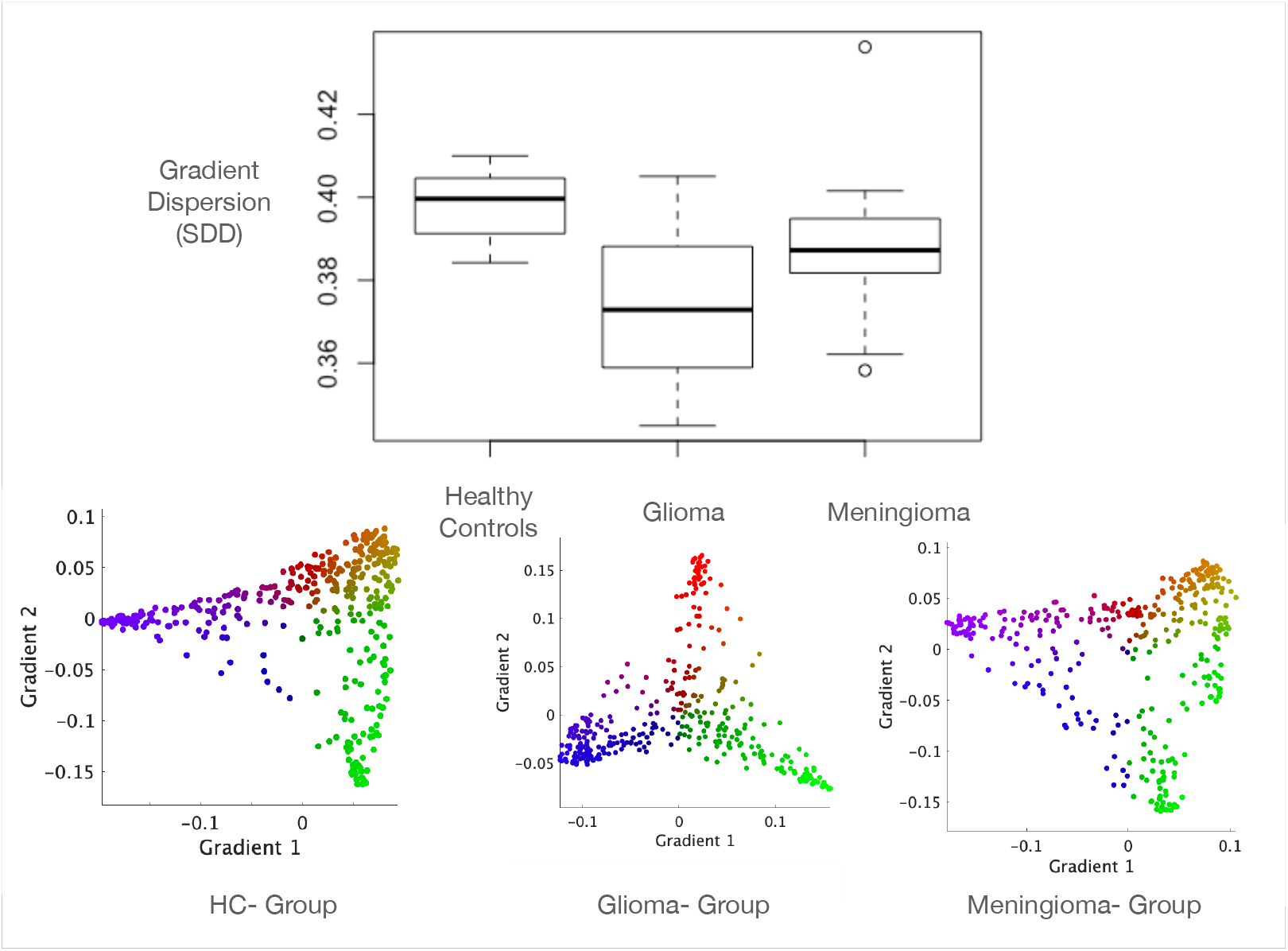
Contrasting Cortical Gradients from Gliomas, Meningiomas, and Healthy Controls. To contrast the glioma cortical gradients, we used this Belgian independent dataset to compare it to meningioma patients and healthy controls^55^. In brief, each subject for each group underwent normalized gradient mapping with SD calculated at the individual level. The top panel of Figure 4 boxplot captures the distribution of dispersion values for each group while the bottom panel illustrates the group-averaged gradient maps. ANOVA revealed a significant difference existed between the three groups (p=0.0121). Comparing gliomas with healthy controls, a significant decrease in gradient dispersion existed (p=0.00795) while there was no significant between healthy controls and meningioma patients (p=0.06).

### Post-operative cortical gradients predict cognitive recovery

A clear stratification emerged when predicting long-term 12-month cognitive outcomes in glioma patients from the SD of cortical gradients at Month 3 (Figure 5). Table S1 summarizes long-term improvers and long-term declines based on neuropsychological testing. Patients experiencing long-term cognitive improvements had an increased gradient dispersion compared to patients who declined long-term and had a lower SD (Figure 5B). This suggests that the cortical hierarchy becomes untethered and increases in dispersion for surgical patients who experience long-term cognitive improvements. Crucially, in patients stratified as Improved or Declined at Month 3, there was no significant difference in pre-surgical tumour volume (p=0.2468, Figure S2), IDH1 mutation status (p= 0.3827), and medical treatment decisions (i.e. chemotherapy or observation, p=0.1763). Lastly, when assessing for gradient stability over time, there was no significant difference between month3 and month 12 gradient dispersion (Figure S6, p=0.09), suggesting there is stability of gradients longer after surgery and the post-operative to month 3 period as a crucial window for rehabilitation.

**Figure 5.**
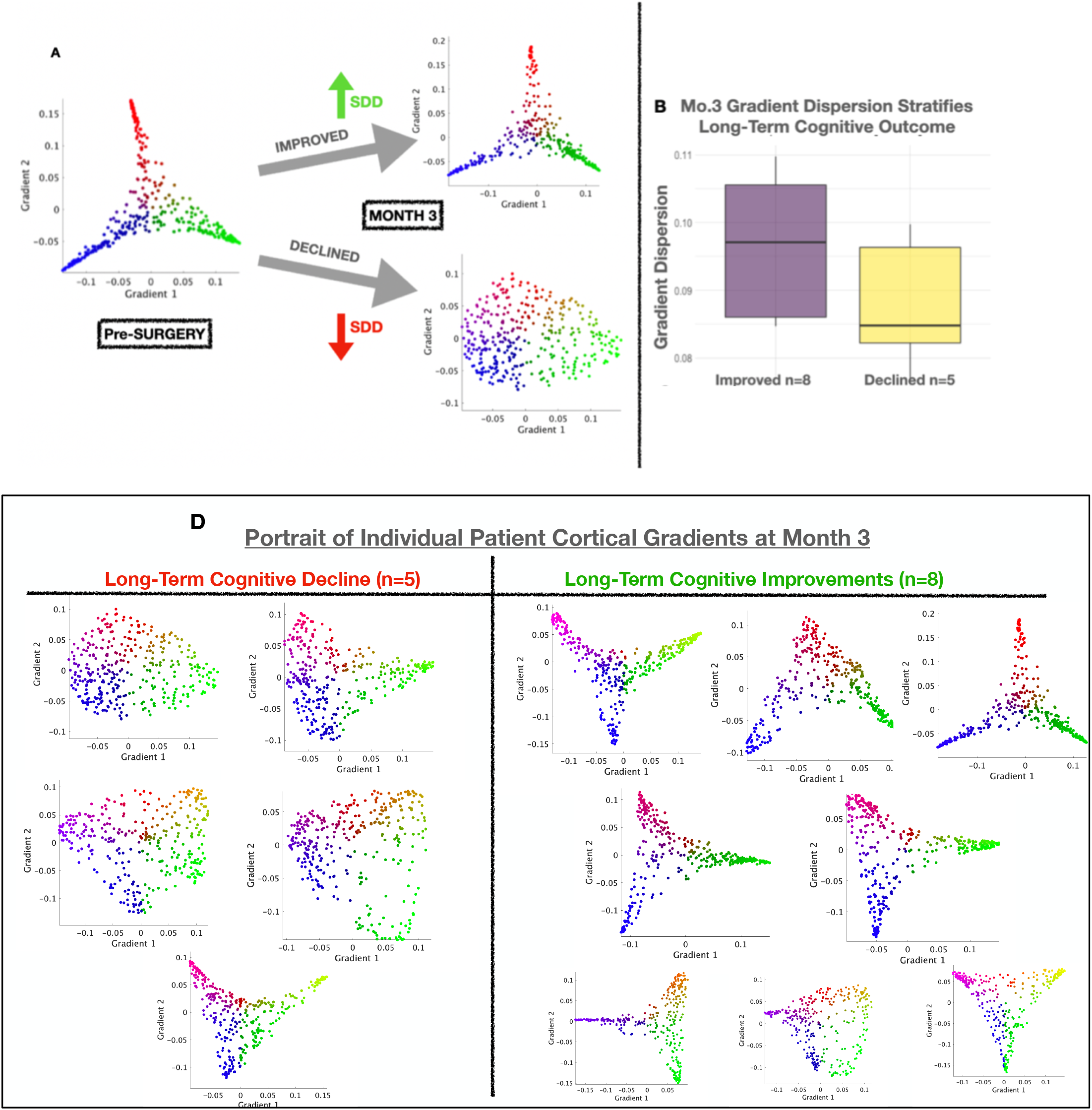
Long-Term Cortical Gradients following Neurosurgery and Rehabilitation. **Top Panel. A)** Stratification of long-term cognitive trajectories based on cortical gradient mapping. By Month 3, post-surgical cortical gradients averaged across patients with subsequent cognitive improvement demonstrate gradient alignment, whereas patients with cognitive decline demonstrated gradient fragmentation. **Right Panel. B)** Patients who cognitively declined had an increased SD compared to patients who improved (Wilcoxon sign ranked test p=0.043). **Bottom Panel:** Month 3 single-subject gradient plots of patients who experienced long-term cognitive decline (bottom-left) and long-term cognitive improvements (bottom-right). Note, only patients with available Month 3 connectomic data were included in this data (n=13).

There was no significant difference between pre-surgical and immediate post-surgical gradient dispersion (p=0.312) Figure S3. Importantly, immediate post-surgical gradient dispersion did not predict long-term cognitive outcomes (r= -0.06, p=0.8266, Figure S5), suggesting a temporal dimension to rehabilitation. There was no significant relationship between pre-surgical tumour volume and pre-surgical gradient dispersion (r=0.113, p=0.666); Figure S4. Finally, spatial autocorrelation analyses between Gradient 1 from healthy controls, glioma, and meningioma patients revealed strong positive correlations between controls and meningioma gradients (r=0.921, p<0.0001), but a strong negative correlation between controls and glioma gradients (r= -0.8549, p<0.0001).

## Discussion

Integrating data from a large normative healthy cohort with a deeply sampled rare neurosurgical cohort, we provide evidence that single-subject gradient can predict executive functioning (EFs) across the lifespan and long-term cognitive outcomes after neurosurgery. By projecting high-dimensional functional MRI data into a low-dimensional, we analyzed glioma-affected brains in a unified space, capture key features of corticocortical functional connectivity to predictive cognitive outcomes. Controlling for age and sex, we found that i) cortical gradient dispersion predicts EFs in healthy individuals, ii) gliomas integrate into functional networks by maintaining cortical gradient shape while reducing dispersion, and iii) neurosurgery induces a bifurcation in cortical gradient trajectories –aligned versus fragmented – that stratifies long-term patient cognitive outcomes.

Irrespective of the grade or location of the tumour, gliomas are infiltrative tumours that integrate with the brain’s parenchyma. Our data reveals a new functional perspective on how gliomas embed themselves within the broader connectome landscape. Specifically, the data suggests that gliomas embed themselves within the cortical hierarchy of functional brain networks by preserving the shape of the manifold (i.e, high levels of similarity in unimodal/sensorimotor and low levels in transmodal/default mode cortical areas) but decreasing overall gradient dispersion compared to controls. This finding supports the prevailing theory that the slow growth of low-grade gliomas (LGGs) allow for minimal disruption of cognitive processes^2,14^. It also builds on recent research showing that gliomas tissue remains functionally active and exerts long-range effects to execute cognitive processes^3,27,37^. Interestingly, no relationship was observed between tumour volume and gradient dispersion in the analysis. Recognizing the present analyses contained a limited neurosurgical sample size and lack of serial presurgical data, one interpretation of the decreased dispersion induced by gliomas is that it insidiously remodels the connectome to accommodate glioma tumours. Our analysis in healthy individuals showed that higher gradient dispersion correlates with reduced executive functioning; thus, the reduced gradient dispersion caused by gliomas may reflect the brain’s functional compensation to maintain communication dynamics and cognitive function despite the presence of glioma. Future studies with larger and more diverse samples could explore how gliomas longitudinally impact the connectome prior to surgery and how different tumour characteristics (i.e, molecular, histological, and imaging features) impact the overall cortical manifold. This glioma-neural integration along the cortical gradient raises critical questions about how brain-wide functional networks interact with glioma tissue and whether specific networks influence tumor growth or inhibition.

At the molecular level, a key question arises: how do gliomas intricately preserve the cortical hierarchy while reducing gradient dispersion? Recent studies suggest potential mechanisms, including increased neuronal excitability^3^ driven by glioma infiltration and the localization of oligodendrocyte precursor cells (OPCs) and microglia in transmodal brain areas^38^. These findings point to the possibility that decreased global cortical gradient dispersion in gliomas may be linked to heightened local expression of OPCs, microglia, and glutamatergic neurotransmission. Thus, consistent with connectome wiring principles^52,53^, the decreased gradient dispersion induced by gliomas may be the result of facilitating more efficient network topology and transitions than alternative more costly topologies. Future research could leverage the Allen Human Brain Atlas (AHBA) and neurotransmitter-receptor density maps to further untangle the relationship between the underlying neurobiology, functional gradients, and glioma behavior^51^.

Our findings highlight that neurosurgical resection, rather than the tumor itself, plays a more significant role in shaping long-term cognitive outcomes. Gradient analyses revealed a distinct bifurcation in patients’ cortical gradients three months post-surgery, whereas immediate post-surgical gradient dispersion did not predict long-term cognitive performance. Patients who showed cognitive improvement over time exhibited the restoration of cortical functional gradients, even in the context of highly altered structural anatomy—offering new evidence for the “tethering hypothesis” of macroscale cortical network organization^39^. Conversely, patients who did not show cognitive improvement retained disrupted cortical gradients with reduced dispersion. From a ‘functional’ perspective within the onco-functional balance framework in surgical neuro^1,14^, glioma resection emerges as a critical factor that alters cortical gradients and triggers the functional plasticity needed for cognitive recovery. Considering the temporal nature of gradient plasticity, early cognitive rehabilitation after surgery could be essential to optimize the restoration of cortical gradients and improve long-term outcomes^41^.

Moving forward, future research could characterize the impact of stereotactic lesioning or electrical stimulation on cortical manifolds. Techniques such as deep brain stimulation, motor cortex stimulation, and focused ultrasound for neurological and psychiatric disorders may offer valuable insights into how cortical gradients are disrupted by targeted, focal interventions^46-48^. These findings could then be contrasted to the results presented in this study on gradient alterations following resective neurosurgery. Additionally, while recent studies have advanced structure-function coupling models of brain connectivity, the mechanisms linking structure-function coupling in cortical gradients remains unclear. By leveraging neurosurgery as a scientific tool, we may better be able to shed light on this relationship.

The present study should be interpreted with acknowledging important methodological considerations. Although some analyses were replicated in an independent dataset, there was participant drop-out at multiple time points. Nonetheless, this remains the only deeply sampled, longitudinal dataset that tracks patients through neurosurgical intervention and recovery. In the absence of comparable datasets in the literature, this study serves as a benchmark for future power calculations. Our dataset was limited to frontotemporal tumors, primarily located in transmodal regions of the cortical hierarchy. Although extremely rare, complementing these present analyses with gliomas in unimodal regions (i.e., motor strip or V1 visual area) would be a valuable avenue for future work. Additionally, these analyses focused exclusively on cortical gradients, excluding subcortical regions, as significant plasticity following glioma resection occurs at the cortical level through horizontal connectivity^39^. For widespread adoption of ‘interventional rehabilitation’ protocols in neuro-oncology^41^, efforts should likely concentrate on modulating cortical areas, which are more accessible and responsive, rather than subcortical regions that may require more invasive procedures^42^.

## Conclusion

This study is the first to demonstrate the methodological utility of gradient mapping in a neurosurgical context. The findings reveal a distinct bifurcation in cortical manifolds between patients with effective and ineffective long-term cognitive rehabilitation. Patients who experienced cognitive improvement after glioma resection showed successful reconstitution of their cortical manifolds, whereas those with long-term deficits exhibited impaired manifold reconstitution. Future research should focus on optimizing surgical planning and designing targeted post-operative rehabilitation programs to minimize cortical manifold disruption and facilitate its recovery during the critical first weeks after surgery.

## Data Availability

All data produced in the present study are available upon reasonable request to the authors

## Supplementary Material

### Supplementary Methods

To replicate our pre-surgical gradient mapping results in our main cohort, we accessed a Belgian brain tumour dataset that was available on OpenNeuro^1^. This dataset contained pre-operative data for 11 glioma patients, 14 meningioma patients, and 11 healthy controls. Specifically, glioma, meningioma, and healthy subject characteristics are tabulated below (Table S1). Table S1 is reproduced from the OpenNeuro dataset that was publicly shared by the authors^1^. Finally, MRI acquisition parameters of patients in this Belgian dataset have been described elsewhere previously^1,2^. The data was pre-processed using the Methods described in the Main text’s Methods.

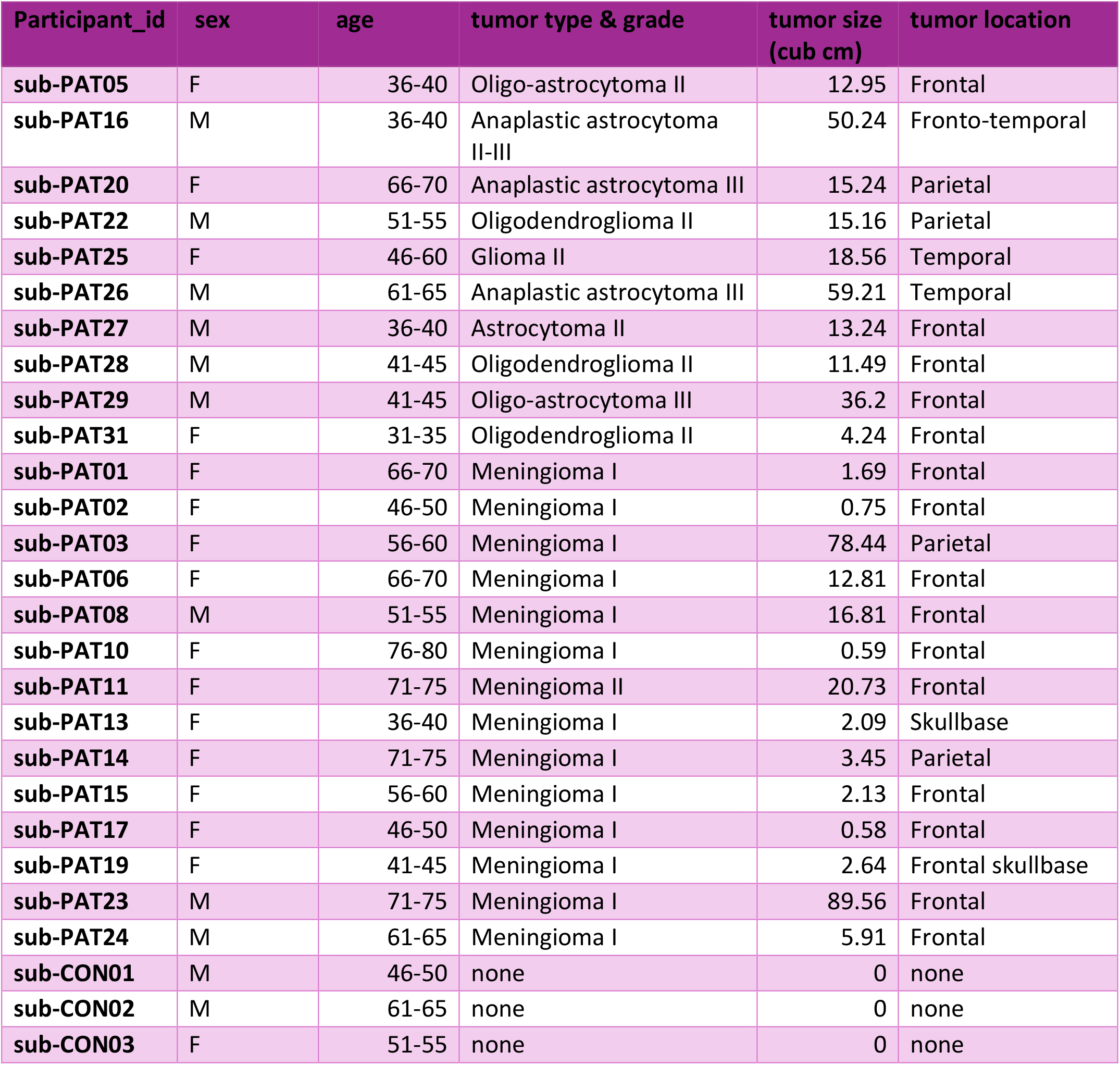
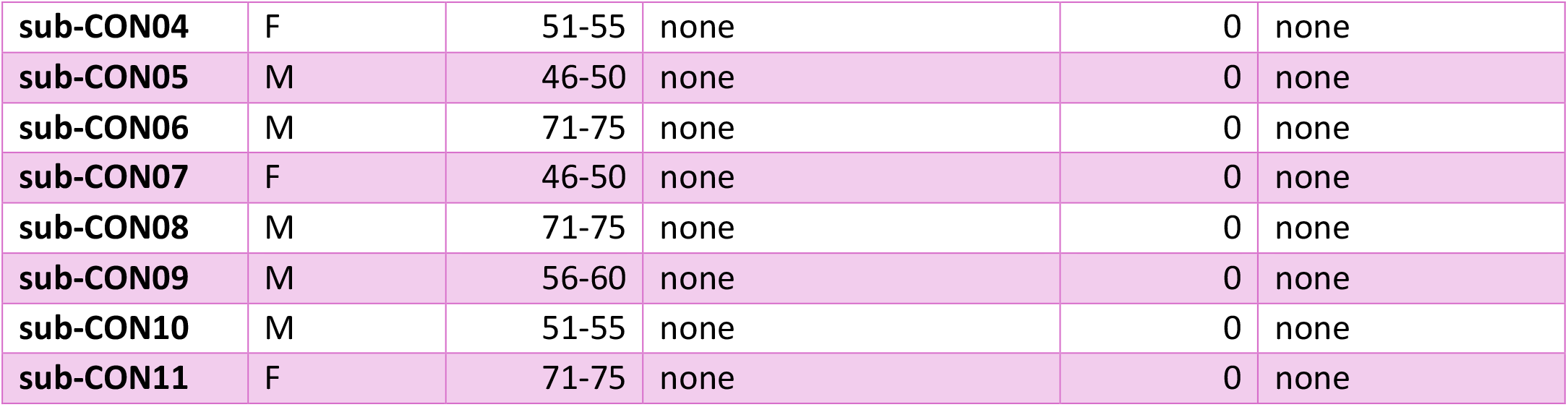

| Participant_id | sex | age | tumor type & grade | tumor size (cub cm) | tumor location |
| --- | --- | --- | --- | --- | --- |
| sub-PAT05 | F | 36-40 | Oligo-astrocytoma II | 12.95 | Frontal |
| sub-PAT16 | M | 36-40 | Anaplastic astrocytoma II-III | 50.24 | Fronto-temporal |
| sub-PAT20 | F | 66-70 | Anaplastic astrocytoma III | 15.24 | Parietal |
| sub-PAT22 | M | 51-55 | Oligodendroglioma II | 15.16 | Parietal |
| sub-PAT25 | F | 46-60 | Glioma II | 18.56 | Temporal |
| sub-PAT26 | M | 61-65 | Anaplastic astrocytoma III | 59.21 | Temporal |
| sub-PAT27 | M | 36-40 | Astrocytoma II | 13.24 | Frontal |
| sub-PAT28 | M | 41-45 | Oligodendroglioma II | 11.49 | Frontal |
| sub-PAT29 | M | 41-45 | Oligo-astrocytoma III | 36.2 | Frontal |
| sub-PAT31 | F | 31-35 | Oligodendroglioma II | 4.24 | Frontal |
| sub-PAT01 | F | 66-70 | Meningioma I | 1.69 | Frontal |
| sub-PAT02 | F | 46-50 | Meningioma I | 0.75 | Frontal |
| sub-PAT03 | F | 56-60 | Meningioma I | 78.44 | Parietal |
| sub-PAT06 | F | 66-70 | Meningioma I | 12.81 | Frontal |
| sub-PAT08 | M | 51-55 | Meningioma I | 16.81 | Frontal |
| sub-PAT10 | F | 76-80 | Meningioma I | 0.59 | Frontal |
| sub-PAT11 | F | 71-75 | Meningioma II | 20.73 | Frontal |
| sub-PAT13 | F | 36-40 | Meningioma I | 2.09 | Skullbase |
| sub-PAT14 | F | 71-75 | Meningioma I | 3.45 | Parietal |
| sub-PAT15 | F | 56-60 | Meningioma I | 2.13 | Frontal |
| sub-PAT17 | F | 46-50 | Meningioma I | 0.58 | Frontal |
| sub-PAT19 | F | 41-45 | Meningioma I | 2.64 | Frontal skullbase |
| sub-PAT23 | M | 71-75 | Meningioma I | 89.56 | Frontal |
| sub-PAT24 | M | 61-65 | Meningioma I | 5.91 | Frontal |
| sub-CON01 | M | 46-50 | none | 0 | none |
| sub-CON02 | M | 61-65 | none | 0 | none |
| sub-CON03 | F | 51-55 | none | 0 | none |
| sub-CON04 | F | 51-55 | none | 0 | none |
| sub-CON05 | M | 46-50 | none | 0 | none |
| sub-CON06 | M | 71-75 | none | 0 | none |
| sub-CON07 | F | 46-50 | none | 0 | none |
| sub-CON08 | M | 71-75 | none | 0 | none |
| sub-CON09 | M | 56-60 | none | 0 | none |
| sub-CON10 | M | 51-55 | none | 0 | none |
| sub-CON11 | F | 71-75 | none | 0 | none |

### Supplementary Results

**Table S1.** Summary of Number of Pre-Surgery Deficits and Post-Surgical Deficits in CAESAR Cohort

| Patient | Pre-Surgery Deficits | Post-Surgical Deficits* | Long-Term Outcome** |
| --- | --- | --- | --- |
| 1 | 1 | 0 | Improved |
| 2 | 0 | N/A | N/A |
| 3 | 1 | 0 | Improved |
| 4 | 1 | 0 | Improved |
| 5 | 2 | 0 | Improved |
| 6 | 1 | 1 | Improved |
| 7 | 0 | 0 | Improved |
| 8 | 2 | 0 | Improved |
| 9 | 1 | 2 | Worse |
| 10 | 1 | 0 | Improved |
| 11 | 0 | 5 | Worse |
| 12 | 4 | 0 | Improved |
| 13 | 0 | 0 | Improved |
| 14 | 0 | 1 | Worse |
| 15 | 2 | 5 | Worse |
| 16 | 0 | 1 | Worse |
| 17 | 0 | 0 | Improved |

\*Post-surgical deficits obtained from latest available follow-up of data.
\*\*Patients who remained the same, pre/post-surgery, were categorized as “improved”

**Figure S1.**
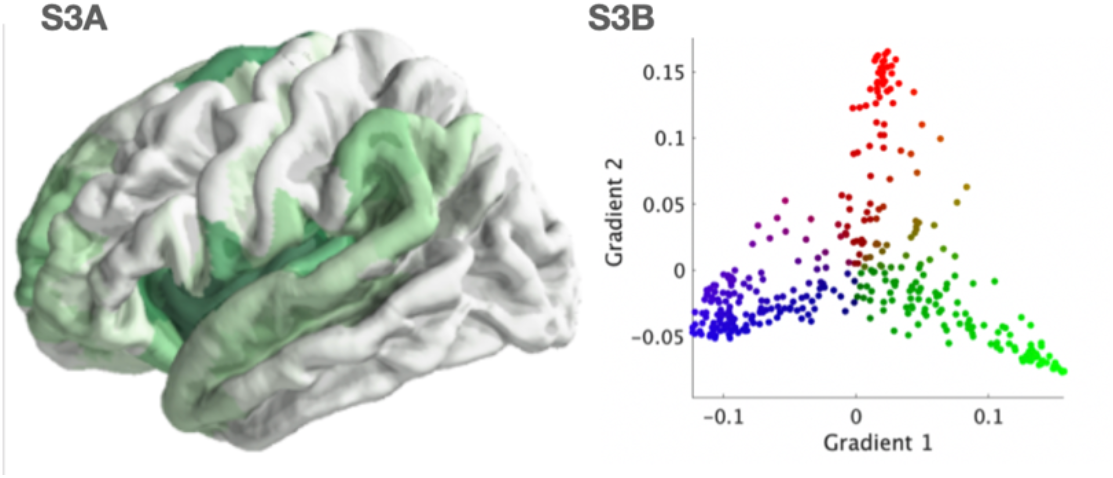
Replication of Glioma Dispersion in Independent Dataset. To replicate our pre-surgical gradient mapping results in our main cohort, we accessed the Aerts Belgian dataset^1^ on OpenNeuro^1^. This dataset contained pre-operative data for 11 glioma patients, 14 meningioma patients, and 11 healthy controls. Figure S3A illustrates that the distribution of gliomas in this Belgian dataset is also frontotemporal while Figure S3B demonstrates the group-level gradient mapping applied to glioma subjects only.

**Figure S2.**
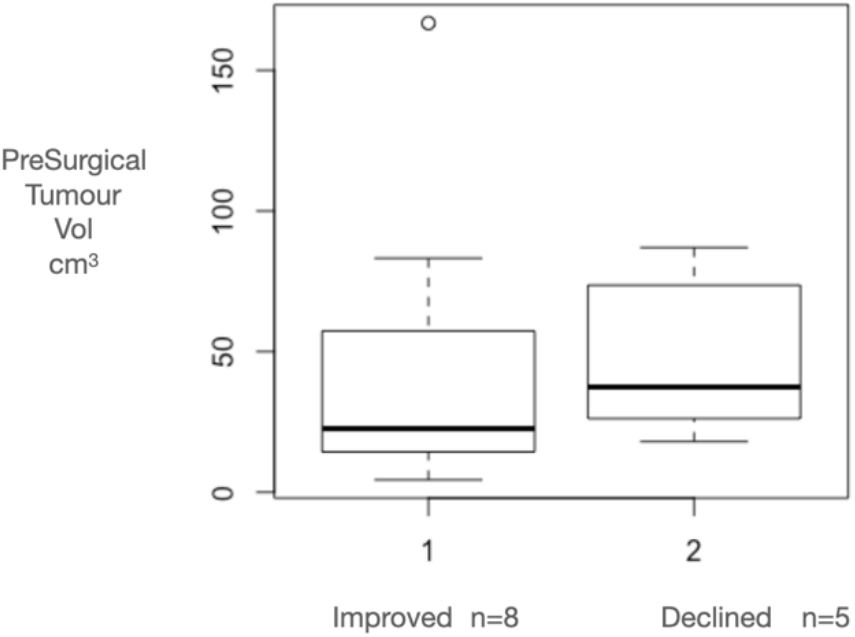
Effect of Pre-Surgical Tumour Volume between patients who improved/declined at Month 3. To ensure that the stratification of long-term cognitive improvements or decline in Figure 4 was not cofounded by the glioma’s volume, we compared pre-operative tumour volumes between both groups. Non-parametric wilcoxon sign ranked test revealed an insiginficant difference (p=0.5303) between both groups. This was also true when removing the outlier in the improved group (p=0.2468).

**Figure S3.**
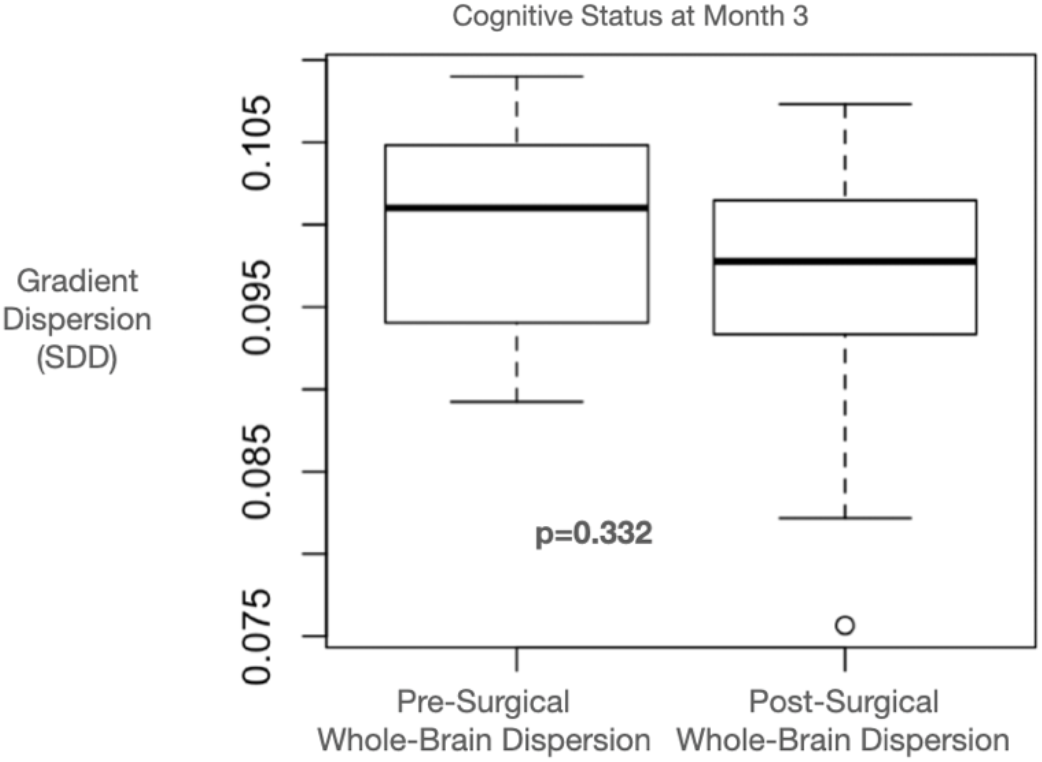
Effect of Cortical Resection on Cortical Gradient Dispersion. To determine if there was an acute effect of cortical neurosurgery on gradient dispersion, we compared pre-surgical and post-surgical gradients across the whole-brain (including tumour pre-surgically, but excluding resection zone post-surgery). There was no significant difference (p=0.332) on non-parametric t-test (Figure S6).

**Figure S4.**
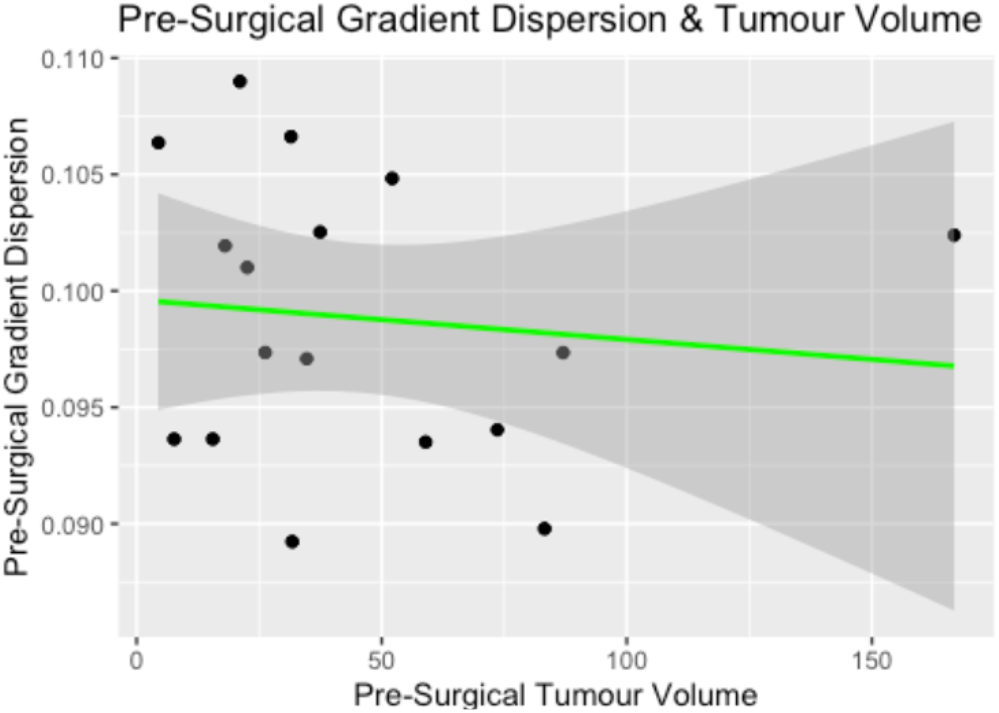
Effect of Pre-Surgical Tumour Volume on Pre-Surgical Gradient Dispersion. To determine if pre-surgical tumour volume affected the pre-surgical gradient dispersion, we conducted a simple linear pearson correlation between both variables. There was no significant relationship between preoperative tumour volume and preoperative gradient dispersion (r=0.1134, p=0.666); Figure S7.

**Figure S5.**
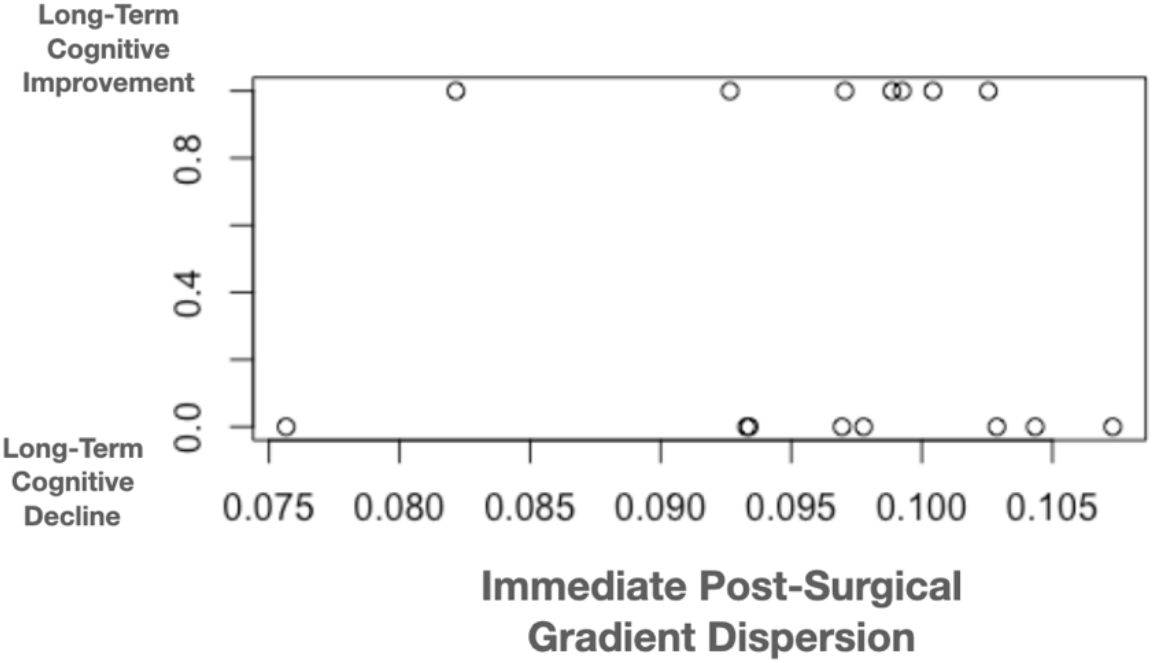
Effect of Immediate Post-Surgical Gradient Dispersion on Long-Term Cognitive Outcomes. To determine if the immediate post-surgical gradient dispersion can predict long-term cognitive outcomes, we performed a spearmann correlation between post-operative dispersion and cognitive outcomes. The analyses revealed a very weak correlation (r= - 0.0618, p=0.8266); Figure S8.

